# Predictors of Physical and Mental Health in Healthcare Teams Working with COVID-19 patients: a scoping review protocol

**DOI:** 10.1101/2020.12.11.20247304

**Authors:** Nelson Aguirre-Duarte, John Øvretveit, Timothy Kenealy

**Affiliations:** Manager & Researcher - High-Reliability Consulting. Honorary academic, School of Population Health, Faculty of Medicine, The University of Auckland, New Zealand; Professor of Health Improvement, Implementation and Evaluation, Medical Management Centre, The Karolinska Institutet, Stockholm, Sweden; Associate Professor of Integrated Care, University of Auckland, New Zealand

**Keywords:** COVID-19, predictors, physical health, mental health, healthcare workers, scoping review

## Abstract

**Introduction:** As a result of the current pandemic (COVID-19), many clinical teams are exposed to stressful situations that may lead to physical and mental issues for clinical staff themselves (we exclude the effects of personal infection with the virus). Recent studies suggest some predictors could depend on context, notably country and the type of the health system.

**Methods and Analysis:** This protocol was follows using the PRISMA-ScR guideline (Preferred Reporting Items for Systematic reviews and Meta-Analyses extension for Scoping Reviews), which was revised and approved by the research team. This study aims to identify factors and evidence of the physical, behavioural and mental consequences of sustained clinical practice in a continuing pandemic. Our research seeks to fill this gap in the literature, and the results may suggest to governments, healthcare authorities and healthcare providers appropriate measures to mitigate risks to healthcare workers during a pandemic response.

**Dissemination and ethics:** The current research design is based on the use of publicly available information and does not require ethical approval. The findings will be disseminated in conferences. Results will be published and additionally shared with relevant local and national authorities.

**Strengths and Limitations of the study:** This will be the first scoping review to identify factors and evidence of the physical, behavioural and mental consequences of sustained clinical practice in a continuing pandemic with health impacts for clinical staff.

The search strategy includes six electronic databases with peer-reviewed literature, as well as a broad range of grey literature sources.

Although this study will not require a quality appraisal, which is consistent with the framework proposed by Arksey and O’Malley, the current study will formally assess the studies quality.

This scoping review study has been registered with Open Science Framework to enhance its transparency.

The search strategy proposed is broad, but the search strategy is limited to articles published in English, Spanish, Portuguese or Italian.

## Introduction

As a result of the current pandemic (COVID-19), many clinical teams are exposed to stressful situations that may lead to physical, mental and behavioural changes affecting their short- and long-term health. Workload and stress may affect safety for staff (and therefore for patients), as well as result in work absences that reduce the number of staff available, compounding the workload and stress for those who remain. Recent studies suggest that these effects are significantly influenced by context, notably country and the type of health system. ^1 2^

There are features of this pandemic which make it difficult to predict effects on staff based on extrapolation from existing knowledge. The pandemic is prolonged, often in volumes that overwhelm local health services, it has proven difficult to predict, control depends on a level of civic cooperation and political support that is not always present, there is a risk of physical harm from infection both to health workers and to their own family and contacts, and an inequitable distribution of effects across multiple domains of society.

This study aims to identify factors and evidence of the physical, behavioural and mental consequences of sustained clinical practice in a continuing pandemic with health impacts for clinical staff. A scoping review methodology was chosen seems the best option as it aims “to examine the extent, range and nature of the research findings in any detail but is a useful way of mapping fields of study where it is difficult to visualise the range of material that might be available.^3^

Our research seeks to identify missing and significant empirical knowledge of use for prioritising and planning interventions as well as methods or theories that can contribute to research and the science base about the effects of sustained emergency situations on clinical staff. The findings are expected to enable governments, healthcare authorities and healthcare providers to take more effective actions to protect the health and well being of staff and patients during a sustained emergency.

## Methods

This protocol was designed using the PRISMA-ScR guideline (Preferred Reporting Items for Systematic reviews and Meta-Analyses extension for Scoping Reviews) ^3 4^

### Stage 1: identifying the research question

We want to identify the influence of the context in those factors (specific populations, socio-demographic characteristics, type of health system, country, within others), also identify factors specific to Māori in New Zealand and other indigenous people internationally, and to identify and map evidence and identify main concepts, theories, sources and knowledge gaps.

### Definitions

#### Frontline workers

healthcare workers responding to the current COVID-19 pandemic, engaged in direct diagnosis, treatment, and care of patients with COVID-19. ^5^ These might be registered professionals such as doctors, nurses and pharmacists, or persons who are not registered professionals but who provide face-to-face care for patients with COVID-19, in healthcare or administrative roles in a healthcare facility or provide healthcare support at home or in residential care.

#### Context

All periods and duration of follow-up are eligible as long as the study deals with the response to the COVID-19 pandemic (last search: 31 October 2020). For this analysis, studies are counted as ‘single country’ if they reported results from only one country in a paper, regardless of their broader affiliation with multi-country research programs. Where papers report results from more than one country date is reported by individual country where possible.

## Research questions

What are the factors that can be associated as predictors of physical and mental health in healthcare workers during a pandemic response – COVID-19 -?

### Stage 2: identifying relevant studies

Articles published in peer-reviewed journal, published conference proceedings, and pre-peer review web publications are potentially eligible. Author NA will conduct literature searches of electronic bibliographic databases in Cochrane Library, EMBASE, Google Scholar, MEDLINE(Ovid), ProQuest, PubMed and Scopus.

The search strategy will be peer-reviewed by author TK using the PRESS (Peer Review of Electronic Search Strategies) checklist. ^6^ A grey literature search will be conducted using Google Scholar search on this topic to consider the first 600 unique references. ^7^ The research strategy incorporates controlled vocabulary and keywords (e.g., (((predictor and “mental health” and physical and “healthcare worker”) or stress or sleep or depression or anxiety) and COVID-19).

### Stage 3: Study Selection

#### Inclusion Criteria

- It is written in English, Spanish, Portuguese or Italian.
- The study will include observational and interventional studies.
- Includes original quantitative data.
- The study subjects and the setting are described in detail
- Healthcare workers (defined above)
- The study examines factors associated with physical and mental health issues in healthcare workers responding to the COVID-19 pandemic.
- Confounding factors are identified and statistical strategies are dealing with them.
- Joanna Briggs Institute methodological guidance for Scoping Reviews index 7 and 8.

### Exclusion Criteria

- Review papers, commentaries, editorials and other publication forms without primary data.
- Scoping reviews and systematic reviews
- The timeframe of the study is not stated (e.g., no detail on when the data was collected).

The search results will be imported from the databases into the online platform RAYYAN QRCI (https://rayyan.qcri.org/). ^8^ Titles and abstracts will be screened for eligibility by two independent reviewers (NA and TK). Studies that meet the inclusion criteria, or if it is unclear whether the research meets the inclusion criteria, will be reviewed in full text. Any disagreement in study selection will be resolved by consensus or by consulting a third reviewer (JO). Reasons for excluding references at the full-text assessment stage of the screening process will be documented and reported in a PRISMA flow diagram. ^9^

### Stage 4 charting data

Two review authors (NA & TK) will independently enter the data from the eligible studies into a custom-designed data extraction form. Disagreements will be resolved by discussion; if consensus cannot be reached, a third author (JO) will review the study and arbitrate.

Data charted will include author, year of publication, sample size, inclusion and exclusion criteria, type of healthcare worker, demographic data (age, gender), study scope (e.g., national or regional, single or multi-country), purpose, country of origin, objectives, methods, situational analysis tool characteristics and use, the inclusion of equity in study design and study limitations. We will be flexible in accommodating additional categories that may emerge during the actual review process, which can aid in comprehensively answering the research question. The data charting domains and subdomains are described in Table 1.

**Table 1.**
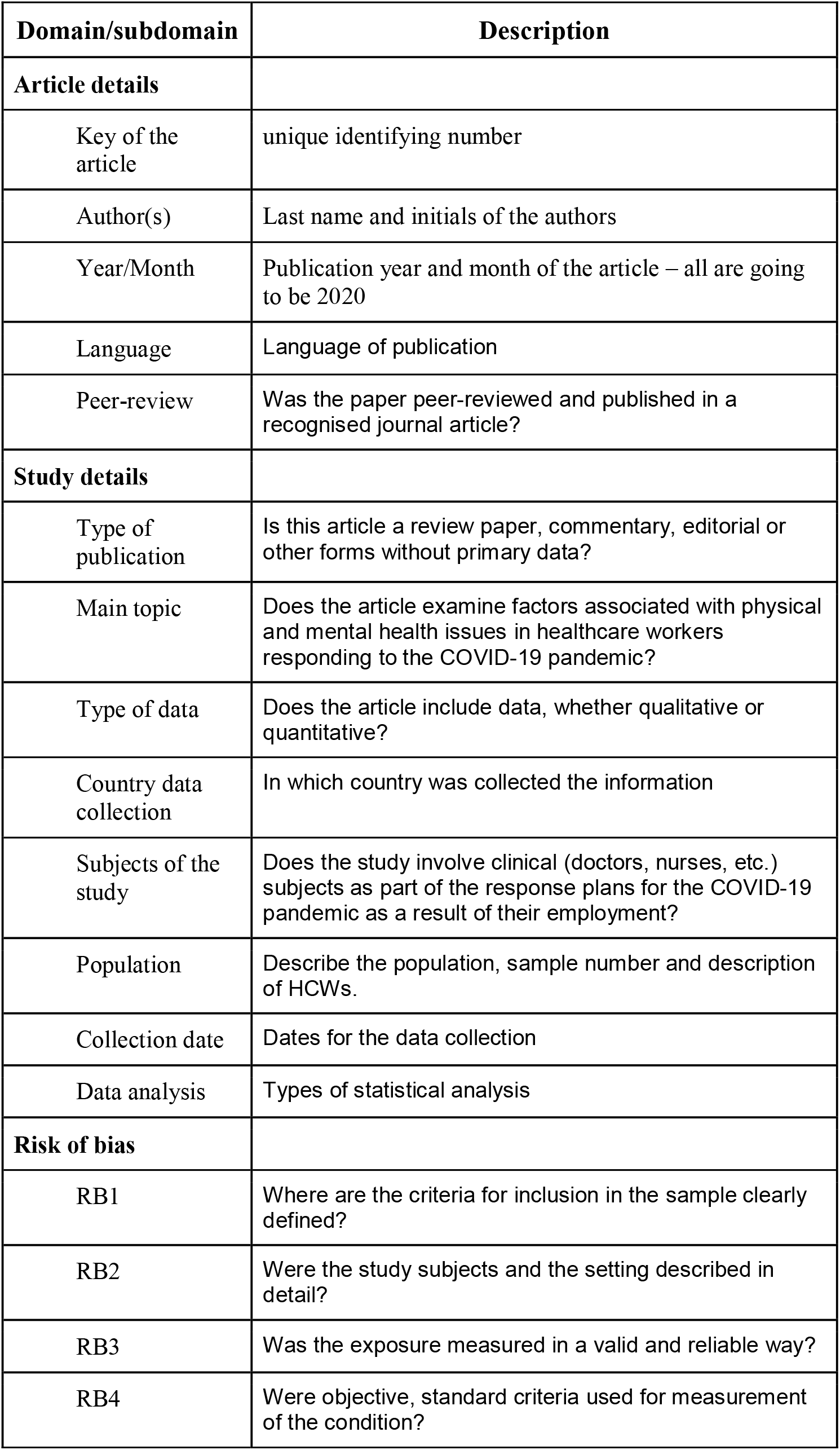
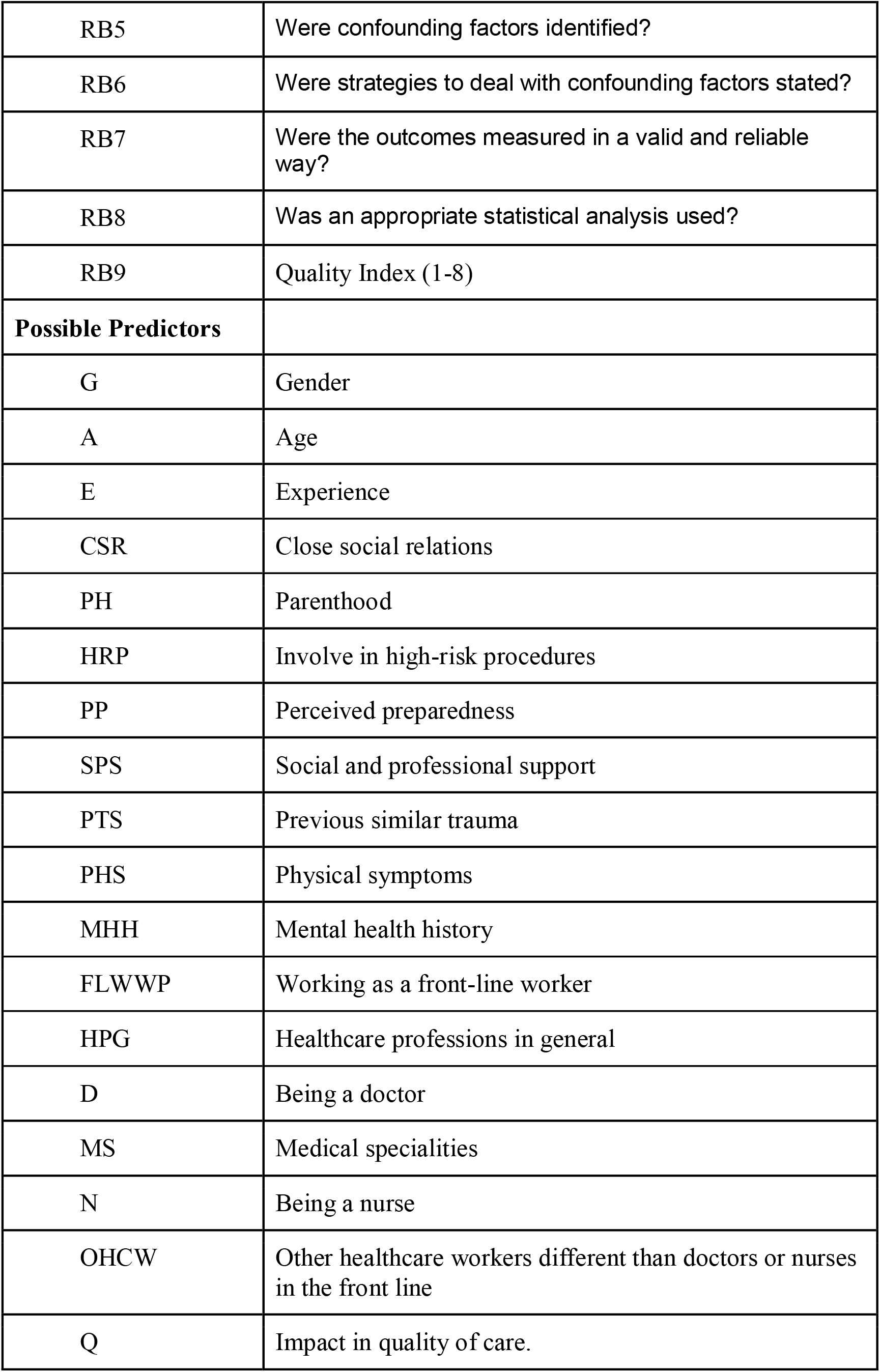
Data charting domains and description of subdomains.

| **Domain/subdomain** | **Description** |
| --- | --- |
| **Article details** |  |
| Key of the article | unique identifying number |
| Author(s) | Last name and initials of the authors |
| Year/Month | Publication year and month of the article – all are going to be 2020 |
| Language | Language of publication |
| Peer-review | Was the paper peer-reviewed and published in a recognised journal article? |
| **Study details** |  |
| Type of publication | Is this article a review paper, commentary, editorial or other forms without primary data? |
| Main topic | Does the article examine factors associated with physical and mental health issues in healthcare workers responding to the COVID-19 pandemic? |
| Type of data | Does the article include data, whether qualitative or quantitative? |
| Country data collection | In which country was collected the information |
| Subjects of the study | Does the study involve clinical (doctors, nurses, etc.) subjects as part of the response plans for the COVID-19 pandemic as a result of their employment? |
| Population | Describe the population, sample number and description of HCWs. |
| Collection date | Dates for the data collection |
| Data analysis | Types of statistical analysis |
| **Risk of bias** |  |
| RB1 | Where are the criteria for inclusion in the sample clearly defined? |
| RB2 | Were the study subjects and the setting described in detail? |
| RB3 | Was the exposure measured in a valid and reliable way? |
| RB4 | Were objective, standard criteria used for measurement of the condition? |
| RB5 | Were confounding factors identified? |
| RB6 | Were strategies to deal with confounding factors stated? |
| RB7 | Were the outcomes measured in a valid and reliable way? |
| RB8 | Was an appropriate statistical analysis used? |
| RB9 | Quality Index (1-8) |
| **Possible Predictors** |  |
| G | Gender |
| A | Age |
| E | Experience |
| CSR | Close social relations |
| PH | Parenthood |
| HRP | Involve in high-risk procedures |
| PP | Perceived preparedness |
| SPS | Social and professional support |
| PTS | Previous similar trauma |
| PHS | Physical symptoms |
| MHH | Mental health history |
| FLWWP | Working as a front-line worker |
| HPG | Healthcare professions in general |
| D | Being a doctor |
| MS | Medical specialities |
| N | Being a nurse |
| OHCW | Other healthcare workers different than doctors or nurses in the front line |
| Q | Impact in quality of care. |

Some scoping reviews will not conduct a quality appraisal, which is consistent with the framework proposed by Arksey and O’Malley, as well as the Joanna Briggs Institute methodological guidance for Scoping Reviews. However, we assessed the quality of the articles using the Joanna Briggs Institute tool for non-randomised studies, which was particularly useful for the pre-print articles.^10^

### Stage 5: collating, summarising and reporting the results

In this review, we will first present an overview of the characteristics of the included studies; this may consist of information on year of publication, the country where the research was performed, study design, sample size, healthcare worker demographic characteristics (such as age and gender), physical and mental severity of illness score within others. The research protocol and the list of articles with reasons for inclusion/exclusion will be published in a repository in the Centre of Open Science platform. ^11^ A summary of results will be sent to relevant policy-makers and researchers working with the healthcare teams responding to the COVID-19 crisis in the form of a 1-page policy brief. We intend toll present our results at a healthcare conference and publish in a peer-reviewed journal. Team members will use their networks to encourage broad dissemination of results.

### Patient and public involvement

No patients were involved in this study.

## Data Availability

All references are available in the OSF repository.

https://osf.io/bqd6w/

## Ethics

Since the scoping review methodology consists of reviewing and collecting data from publicly available literature, this scoping review does not require ethics approval.

## Funding sources/sponsors

This study is not funded.

## Competing interest

None declared

## Patient consent for publication

No required

## Notes

### Competing Interest Statement

The authors have declared no competing interest.

### Author Declarations

No ethics application is required for this study.

